# Pleiotropic and sex-specific genetic architecture of circulating metabolic markers

**DOI:** 10.1101/2024.07.30.24311254

**Authors:** Dennis van der Meer, Zillur Rahman, Aigar Ottas, Pravesh Parekh, Gleda Kutrolli, Sara E. Stinson, Maria Koromina, Jaroslav Rokicki, Ida E. Sønderby, Nadine Parker, Markos Tesfaye, Guy Hindley, Linn N. Rødevand, Elise Koch, Estonian Biobank Research Team, Nils Eiel Steen, Jens Petter Berg, Kevin S. O’Connell, Olav B. Smeland, Oleksandr Frei, Anders M. Dale, Srdjan Djurovic, Kelli Lehto, Maris Alver, Lili Milani, Alexey A. Shadrin, Ole A. Andreassen

**Author notes:** Corresponding authors &. Address: Kirkeveien 166, 0450 Oslo, Norway.

## Abstract

**Background:** Metabolites in plasma form biosignatures of a range of common complex human diseases. Mapping the genetic architecture and discovering variants with pleiotropic effects across metabolites can reveal underlying mechanisms and potential targets for personalized interventions.

**Methods:** We performed univariate and multivariate genome-wide association studies (GWAS) on the Nightingale panel of 249 circulating plasma metabolic markers, across 207,836 White British UK Biobank participants (mean age 57.4 years, 53.7% female), with replication conducted across 27,509 UK Biobank participants with different ancestries, and 92,661 Estonian Biobank participants (mean age 50.9 years, 65.7% female). We investigated rare variation through whole exome sequencing gene burden tests, quantified genetic architectures through Gaussian mixture modelling, analysed the causal role of body mass index (BMI) through Mendelian randomization, and performed genome-wide interaction analyses with sex.

**Results:** We discovered 14,837 loci (497 unique), with shared and distinct effects on cardiometabolic traits, with high replication rates across populations. The loci explained over 70% of genetic variance for fatty acids. Findings from common and rare variant gene tests converged on lipid homeostasis pathways. There was strong evidence for causal effects of BMI on cholesterol and amino acid levels. We discovered 31 loci interacting with sex, which mapped to genes involved in cholesterol processing, and to cardiometabolic conditions with sex differences in prevalence.

**Discussion:** The findings offer new insights into the genetic architecture of circulating metabolites, revealing novel loci and plausible sex-specific molecular mechanisms of lipid metabolism. This improved understanding of the molecular biology of metabolism lays a foundation for personalized prevention and treatment strategies.

---

Small molecules abundant in plasma, including lipoproteins, fatty acids, amino acids, and ketone bodies, are part of metabolic processes essential for human health. Reliable quantification of absolute concentrations of metabolites can now be achieved through high-throughput nuclear magnetic resonance (NMR) spectroscopy.^1^ Metabolomics data in large population samples such as the UK Biobank (UKB), coupled to national health records, has allowed researchers to identify numerous associations between patterns of metabolite concentrations and a wide range of common medical conditions.^2^ These metabolites hold potential for precision medicine as they have been shown to predict long term outcomes,^3^ and could aid in combatting key public health issues, including the adverse effects of the worldwide obesity epidemic.^4^

Charting the pleiotropic genetic architecture of metabolic biomarkers, through the effects of common and rare variants, is key to understanding interindividual differences in metabolic processes. Genome-wide association studies (GWAS) of metabolomics data have confirmed there is a substantial genetic component to these metabolite concentrations and have identified dozens of common variants associated with individual metabolites.^5^ The sets of metabolites included in metabolomics panels are strongly genetically correlated to each other;^6^ joint analysis through a multivariate approach may aid in discovery of variants with widespread effects by leveraging shared genetic signal across the metabolites.^7^ Additionally, characterizing the influence of rare variants on metabolites through whole exome sequencing (WES) data complements GWAS efforts, as rare variants are likely to be particularly impactful and point towards promising drug targets.^8,9^

Obesity and sex are likely important moderators of the relation between an individual’s genetic make-up and metabolic health. As obesity and its downstream medical conditions co-occur with changes in metabolite concentrations;^10^ disentangling the causal role of obesity in determining these levels can aid in devising treatment strategies. Biological sex is a further important determinant of metabolic activity,^11^ yet there is little knowledge about sex-dependent genetic influences. Males and females differ substantially in basal metabolic activity, as well as in their propensity to develop prominent metabolic conditions, such as obesity, coronary artery disease (CAD) and type 2 diabetes (T2D).^12^ Previous studies have shown that there is a genetic basis for sex differences in metabolism, beyond the impact of gonadal hormones.^13^

Here, we take advantage of the latest generation of targeted metabolomics technology available in the UKB and Estonian Biobank (EstBB), to perform the largest GWAS to date of circulating metabolic traits, leveraging NMR spectroscopy data from over 300,000 individuals. We employ a multivariate approach to boost discovery of variants with widespread shared effects across metabolites. We additionally perform novel quantification of the global genetic architecture and incorporate WES data, to expand on knowledge about the impact of both common and rare variants. Lastly, we identify widespread sex-specific effects and estimate the influence of obesity (indexed by BMI) to provide a clinically relevant risk scenario as a foundation for precision medicine approaches.

## Results

We conducted GWAS of 249 circulating metabolites from the Nightingale NMR metabolomic platform, charting their shared and specific genetic architectures. This panel encompasses 228 lipids, lipoproteins and fatty acids, and 21 non-lipids, including amino acids, ketone bodies, fluid balance, glycolysis- and inflammation-related metabolites. See Supplementary Table 1 for an overview of these circulating metabolites, their categories, and sample sizes. For the main analyses we used data from UKB, including 207,836 White British participants, with a mean age of 57.4 years (standard deviation (SD) 8.0 years), 53.7% female. Additionally, there were data on 27,509 non-White British UKB participants, with a mean age of 54.5 years (SD=8.4 years), 54.3% female. From EstBB, we included 92,661 unrelated White European participants, with a mean age of 50.9 years (SD=16.2 years), 65.7% female, which we used to test for generalization of the discovered loci across different populations. For each of these subsets, identical analyses were carried out, covarying for age, sex, and the first twenty genetic principal components to control for population stratification.^14^

### Global genetic architecture

We first determined the SNP-based heritability, h^2^, of the metabolites through LD score regression (LDSC). We also estimated the polygenicity and average magnitude of non-null effects (‘discoverability’) by fitting a Gaussian mixture model of null and non-null effects to the univariate GWAS summary statistics using MiXeR.^15,16^ Overall, the output showed that the metabolites vary widely in their global genetic architecture; h^2^ ranged from .02 (acetoacetate, standard error (SE)=.002) to .21 (triglycerides to total lipids in very large HDL, SE=.001), all p-values < 1.1*10^−21^. Polygenicity estimates spread across two orders of magnitude, from oligogenic (phenylalanine with an estimated 17 causal variants) to moderately polygenic (creatinine with an estimated 1529 causal variants). Similarly, discoverability ranged from 1.2*10^−4^ (lactate) to 2.4*10^−3^ (omega-3%). Heritability was driven by polygenicity (Spearman correlation, ρ, =.56, p<1*10^−16^), not by discoverability (ρ=-.11, p=.08), and there was a strong negative relation between polygenicity and discoverability (ρ=-.81, p<1*10^−16^), similar to other biological measures.^17^ **Figure 1** depicts the estimated proportion of h^2^ explained by genome-wide significant variants as a function of sample size, for 37 metabolites validated for clinical use.^1^ This shows that, for some metabolic markers, the GWAS is approaching saturation; over 70% of genetic variance is explained by discovered variants at the current sample size for omega-3 and omega-6 fatty acid concentrations. These results further showed that the genetic architecture varies by metabolite category; lipid levels have a relatively simple architecture while levels of glycolysis- and fluid balance-related metabolites are more complex. All estimates, for each of the 249 metabolites, are listed in Supplementary Table 1.

**Figure 1.**
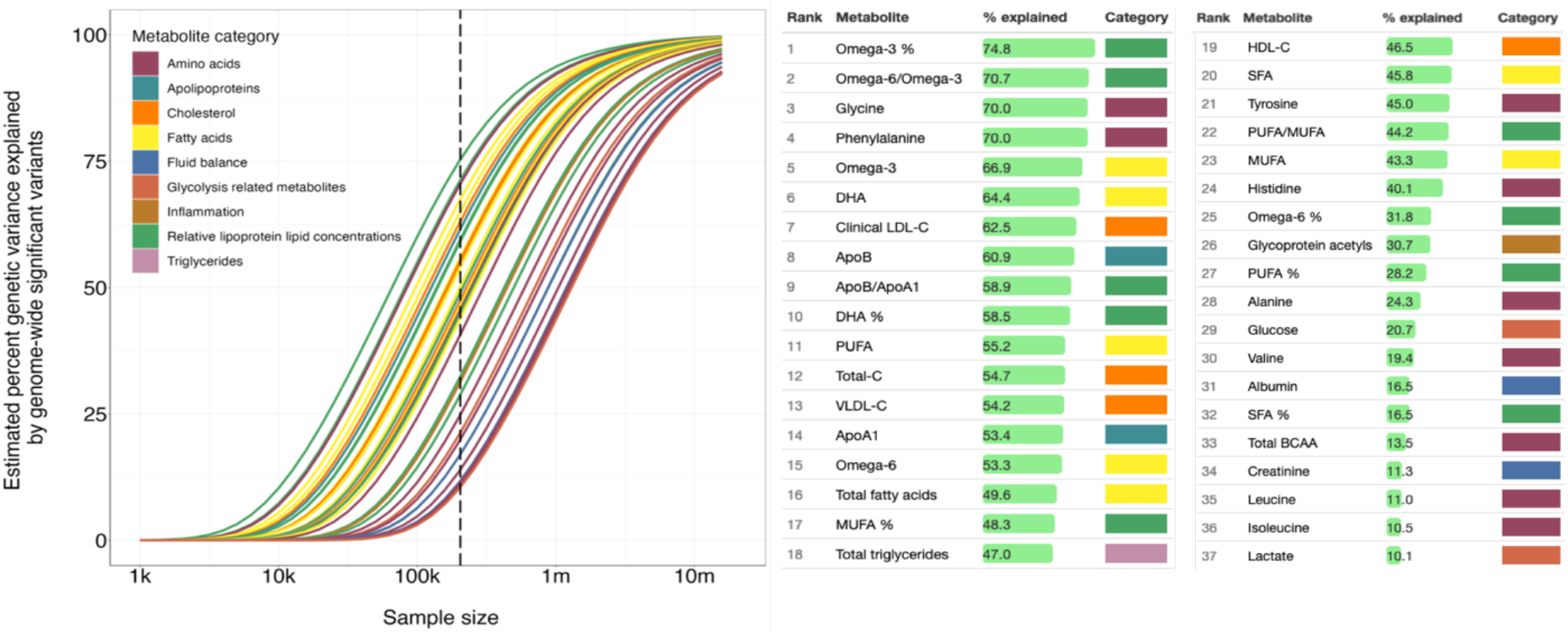
Required sample sizes to uncover the genetic architecture of metabolite concentrations. Illustration of the relationship between genetic variance explained by genome-wide significant hits (y-axis) and sample size (x-axis) for 37 clinically validated metabolites, ordered from most to least explained variance in the table next to the plot. Colours indicate metabolite category, as outlined in the legend. The vertical dashed line marks the current sample size.

### Univariate GWAS

We estimated the effective number of independent traits in our analyses to be 96, based on matrix spectral decomposition^18^ of the phenotypic correlation between all 249 metabolite concentrations. We therefore set the univariate GWAS significance threshold at p=5*10^−8^/96=5.2*10^−10^. The GWAS of all individual 249 metabolites revealed 497 unique loci loci surpassing the significance threshold. There was a median of 61 loci discovered per metabolite (range 8 to 97). When aggregated, there was a total of 14,873 unique loci, as shown in **Figure 2**, suggesting high numbers of shared genetic variants across the metabolites. In line with the differences in estimated discoverability, the number of loci discovered for lipid measures was more than double that for non-lipid measures, as displayed in Supplementary Figure 1. Supplementary Table 1 further lists the number of significant loci and lead SNPs for each of the 249 metabolites.

**Figure 2.**
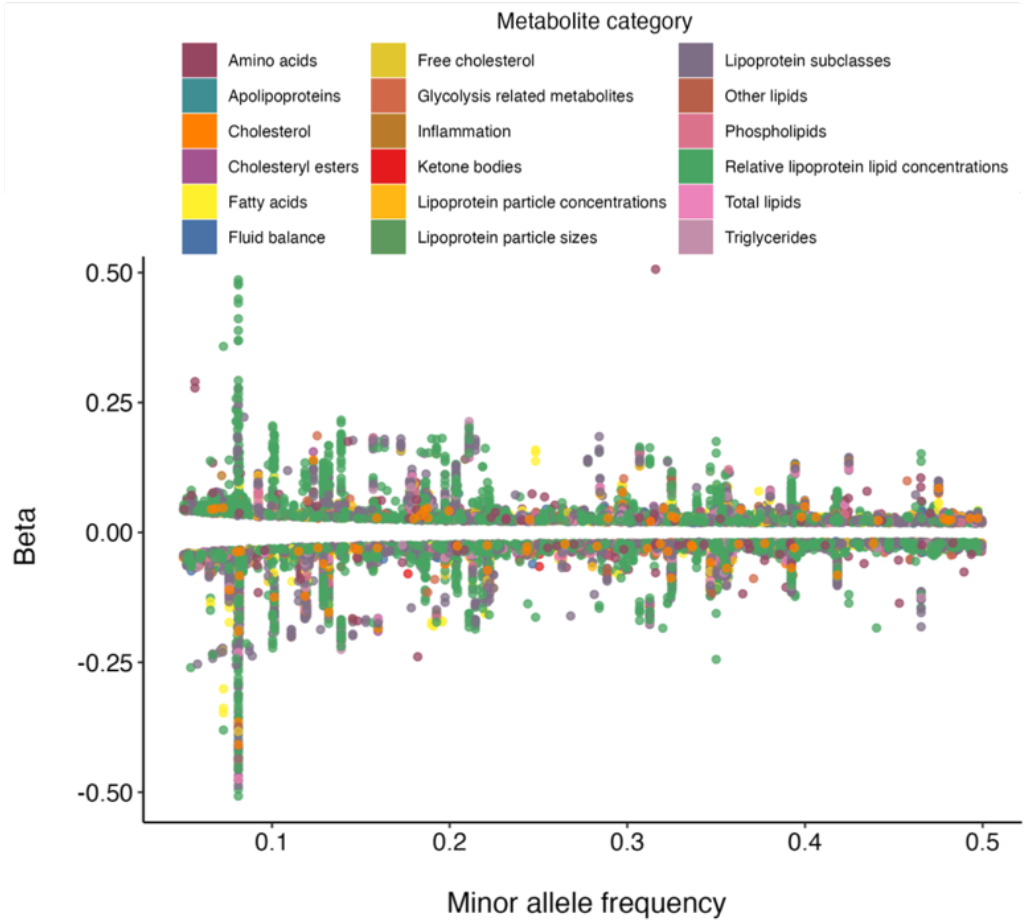
Effect sizes for discovered loci for individual metabolites. Scatterplot displaying the effect sizes (y-axis) of all 14,873 locus lead SNPs identified through univariate GWAS of 249 metabolites, ordered by their minor allele frequency (x-axis) and colour coded by metabolite category.

We checked replication of the locus discovery in the non-White British UKB subset and the White EstBB cohort. We found that 94.9% of all 14,873 discovered locus lead variants showed the same direction of effects in the additional UKB subset (n=27,509 individuals), and 56.4% were nominally significant. For the larger EstBB replication set (n=92,645 individuals), the concordance rate was 99.0% for the 11,386 available locus lead variants, and 92.2% were nominally significant. Thus, our results suggest cross-population generalization of the discovered genetic associations. Supplementary Figure 1 shows the relationship between the number of discovered loci in UKB and replicated loci in EstBB, per metabolite. Supplementary Table 2 lists information on all discovered loci per metabolite, including the replication p-values, and all Manhattan plots are provided in Supplementary Figure 2.

### Multivariate GWAS

Genetic variants are likely to have distributed effects across the metabolites, given these metabolites are correlated components of the same biological system, as also indicated by the univariate GWAS findings. We therefore jointly analysed all measures with the Multivariate Omnibus Statistical Test (MOSTest),^7^ which prioritizes the identification of pleiotropic variants by leveraging shared genetic signal across the univariate measures, yielding a multivariate association with each genetic variant.

For the primary sample, MOSTest revealed 12,216 independent significant SNPs and 2,690 lead SNPs across all metabolites, for a total of 534 loci covering 8.3% of the genome, see **Figure 3a**. The lead SNPs of 96 of these loci did not show genome-wide significant effects on any of the individual metabolites, i.e. they were detected only through MOSTest due to their distributed signal across the metabolites. Supplementary Figure 3 summarizes the significance of the locus lead SNPs across all metabolites, illustrating the pervasive pleiotropy of most discovered variants. Indeed, 48 of these SNPs showed a genome-wide significant association with more than 100 metabolites, as summarized in Supplementary Table 3.

**Figure 3.**
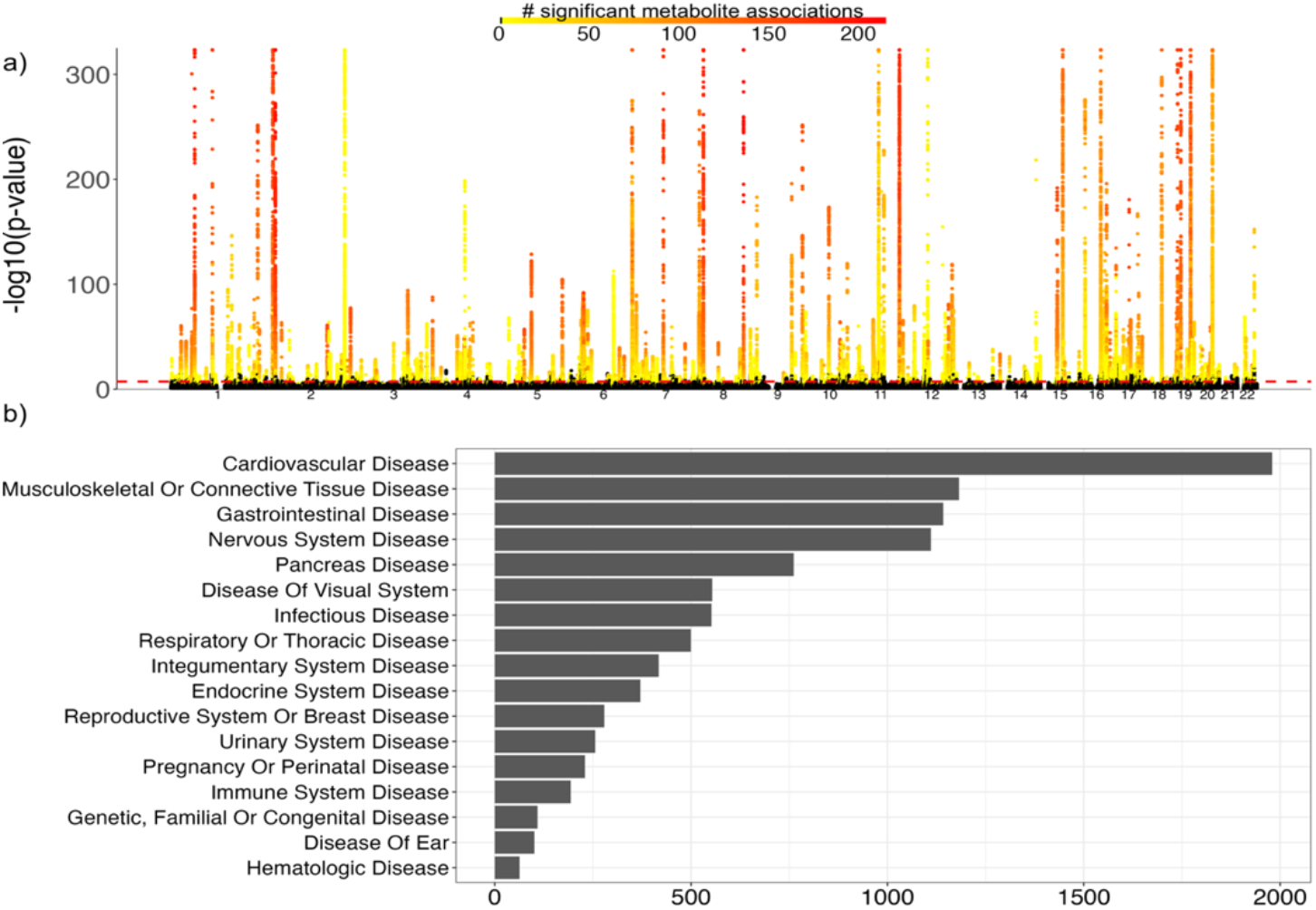
Discovery of pleiotropic variants and their relationship to disease. **a)** Manhattan plot of the output of the multivariate GWAS (MOSTest) on all 249 metabolites, with the observed -log10(p) for each variant shown on the y-axis. The x-axis shows the relative genomic location, grouped by chromosome, and the red dashed line indicates the genome-wide significance threshold of 5×10^−8^. The colour coding represents the number of genome-wide significant associations of each variant with metabolites at the univariate level, ranging from 0 (in black) to 214 (in red), illustrating the extent of pleiotropy. **b)** Bar plot of findings from the phenome-wide association study, coupling the 534 MOSTest-identified lead SNPs to published GWAS of medical conditions. On the x-axis are the number of significant associations, and on the y-axis the categories of diseases as compiled by OpenTargets.

We further compared the findings to the previously largest GWAS of 233 metabolites of the Nightingale metabolomics panel, which identified 276 genomic regions across 136,016 participants and did not include UKB in the discovery sample.^19^ This comparison showed that 274 of these 276 regions overlap with the MOSTest-discovered loci, while MOSTest uncovered another 260 loci not reported in this previous GWAS.

We performed phenome-wide association studies (pheWAS) of each of the 534 loci identified through MOSTest, querying GWAS Catalog and FinnGen GWAS summary statistics through the ‘otargen’ R package, leveraging the OpenTargets ‘diseases’ categorization in order to determine clinical relevance.^20^ There were a total of 9,816 significant associations (p<.005), with 1545 reported traits across 1,621 studies. The results, fully listed in Supplementary Table 4, are summarized in **Figure 3b**. This shows that many of the discovered variants are associated with cardiovascular diseases, as expected. Notably, the nervous system disease category (encompassing psychiatric and neurological diseases) ranks high in this list, among more outright cardiometabolic conditions, fitting with mounting evidence of the importance of metabolic dysfunction in these conditions.^21^

### Gene-based analyses

To gain biological insights, we employed several complementary approaches to gene identification. First, we applied a combination of PolyFun and FINEMAP, Bayesian fine-mapping procedures bundled in the SAFFARI pipeline,^22^ to each of the univariate GWAS summary statistics, retaining 2,625 variants with a posterior probability >.95 as a credible set. We then mapped these variants to 2,493 protein-coding genes using OpenTargets.^23^ Supplementary Table 5 lists the results from fine-mapping in more details, including all mapped genes and their coupling to individual metabolites.

Next, we ran SKAT-O^24^ gene burden tests on WES data restricted to intragenic variants with MAF <.005, to characterize the impact of rare exonic variation on metabolites. There were 335 protein-coding genes with a multiple comparison-corrected significant burden (p<.05/(96 independent traits * 17,849 total protein-coding genes), see Supplementary Table 6.

**Figure 4** lists the top genes identified through burden tests, based on the number of associations with individual metabolites, split by metabolite category. This showcases the widespread impact of apolipoprotein genes, well-known for their association with obesity and Alzheimer’s disease, on these metabolites. We used the DGIdb (v5.0.6)^25^ to identify drug-gene interactions among the 338 genes, showing that 128 genes were interacting with a total number of 1,244 drugs. Many of these are anti-inflammatory, anti-hypercholesterolaemic, or anti-hypertensive, in addition to multiple anti-depressants and anti-psychotics. We performed GSEA to test if the 338 genes were enriched for drug target genes of specific drugs, showing significant (FDR<0.05) enrichment for targets of nine drugs. These include three lipid-lowering agents, three antibacterial drugs, a drug used to treat leukemia, an anticonvulsant, and a platelet aggregation inhibitor (Supplementary Table 7).

**Figure 4.**
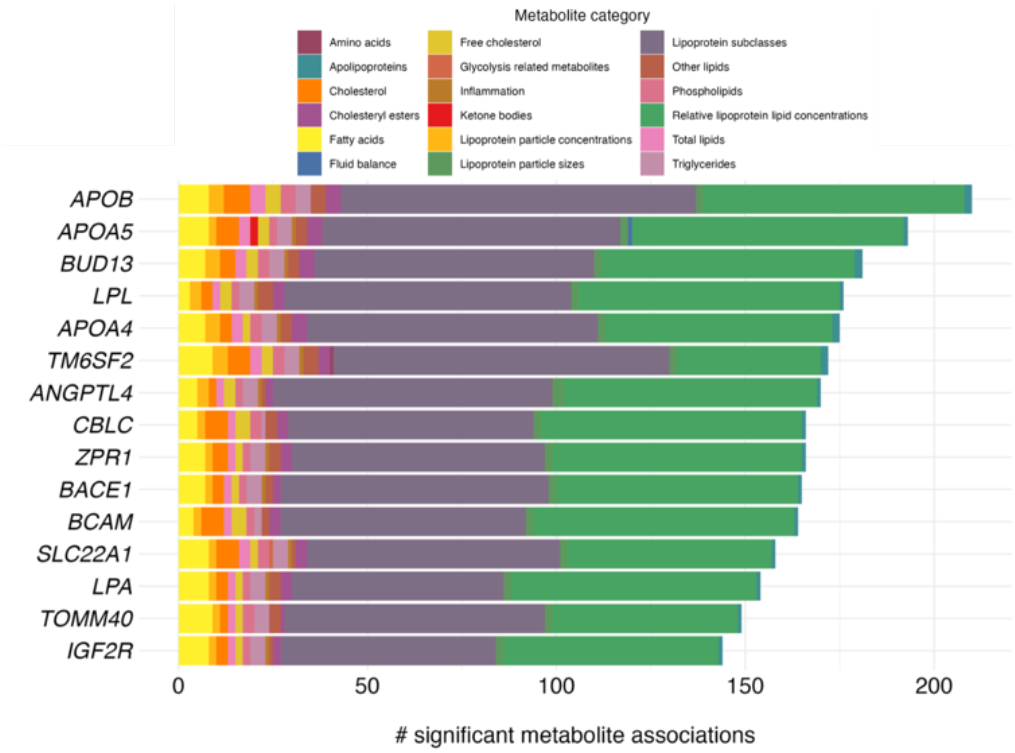
Genes most widely associated with metabolites based on rare variants. Stacked bar plot showing the number of significant associations (x-axis) with genes identified through whole-exome sequencing-based gene burden tests (y-axis), coloured by metabolite category.

Next, we applied MAGMA to each of the univariate GWAS summary statistics, aggregating across all common variants within the same 17,849 genes. Tests of tissue-specificity, covarying for mean expression across all tissues, revealed differential expression in the liver for nearly all metabolites (243 out of 249), in line with its central role in metabolism of both lipids and amino acids. Differential expression of gene-sets in other tissues was more specific to a metabolite category, as can be seen for the spleen, summarized in **Figure 5a**. Competitive gene-set analysis for each individual metabolite GWAS, testing for 7,522 Gene Ontology (GO) biological processes, primarily uncovered associations with lipoprotein particle modification, organization, and homeostasis, see **Figure 5b**. Supplementary Table 8 contains the complete results of the gene-set analyses for each individual metabolite.

**Figure 5.**
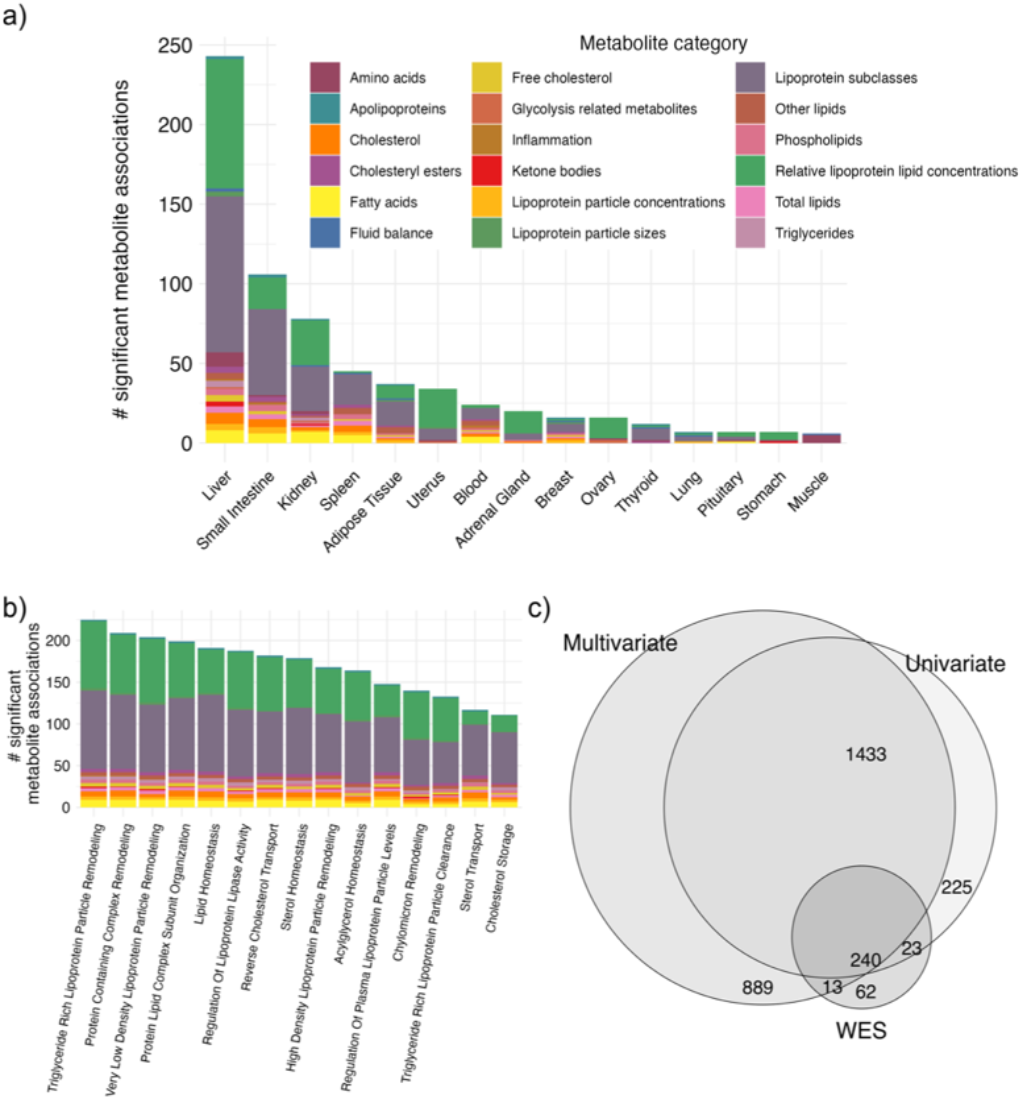
Functional annotation of gene-based tests. **a)** Stacked bar plot summarizing the output of tests of tissue-specific gene expression, with the top 15 tissues on the x-axis. **b)** Competitive gene-set analysis of Gene Ontology biological processes, with top 15 pathways listed on the x-axis. For both plots, the number of significant associations with metabolites is shown on the y-axis and the colours indicate metabolite categories. **c)** Venn diagram of the number of genes identified through gene-based tests of the multivariate GWAS (MOSTest), univariate GWAS and rare variant WES data.

We further applied MAGMA to the multivariate GWAS summary statistics as well, identifying particularly pleiotropic genes. This allowed us to compare the gene-level association of pleiotropic, common and rare variation with the metabolites. We identified 2,590 multiple-comparisons-corrected significant protein-coding genes through the multivariate approach compared to 1,921 through the univariate approach and 335 through the WES burden tests. See **Figure 5c** for overlapping and unique components of the identified sets of genes through these different approaches. Supplementary Table 9 lists all identified genes per gene-based test. The unique and shared sets of genes were coupled to the GWAS Catalog through hypergeometric tests, with results summarized in Supplementary Table 10.

### Analyses of sex and BMI

Given sex and BMI have been associated with substantial interindividual variation in metabolic activity,^10,11^ we next investigated the phenotypic and genetic relation of these individual determinants with the metabolites. First, we conducted linear regression analyses, regressing each metabolite onto sex, BMI, sex*BMI, and age. These models produced highly significant associations with sex, BMI, and their interaction across nearly all metabolites, as summarized in Supplementary Table 11. Notably, there was a very high correlation between the coefficients of sex and BMI (r=.87, p=4.1*10^−79^), indicating that these factors share mechanisms that in turn impact metabolites. This underlines the need for sex-specific research into metabolic health.

### Sex-specific genetic influences

We first ran univariate GWAS within both sexes separately, to compare the overall genetic architecture between men and women. Through paired t-tests applied to sex-specific LDSC heritability estimates, we found that the mean h^2^ was significantly higher for women than for men (h^2^=.148 vs. .132, t=12.8, p<1*10^−16^). Men’s h^2^ was still higher than that of the overall GWAS (h^2^=.132 vs. .128, t=7.5, p=9*10^−13^), suggesting heritability estimates may be lowered by combining two subsamples (men and women) with differing genetic influences. We further calculated genetic correlations between the two sets of sex-specific GWAS and found that these ranged between .85 and 1. While these correlations were high, the majority differed significantly from 1, as reported in Supplementary Table 12.

Given the identification of sex-specific genetic components through LDSC, we ran multivariate GWAS with an interaction term between sex and each genetic variant, to discover individual variants with sex-specific effects. We found 31 loci with a genome-wide significant interaction effect, see **Figure 6a**. Follow-up in the univariate summary statistics showed that the interaction effects were often present for numerous metabolites, with one interaction effect (rs1065853, *APOE*) being genome-wide significant across 110 metabolites. **Figure 6b** provides an example of univariate cross-over interaction effect between sex and the rs1065853 genetic variant on lipid levels, with no significant main effect of this SNP. **Figure 6c** shows another significant sex*gene variant interaction effect of rs964184 (*ZPR1*), a known risk factor for metabolic syndrome and CAD,^26,27^ which appears to only influence cholesterol levels in females. In total, there were 496 univariate genome-wide significant interactions.

**Figure 6.**
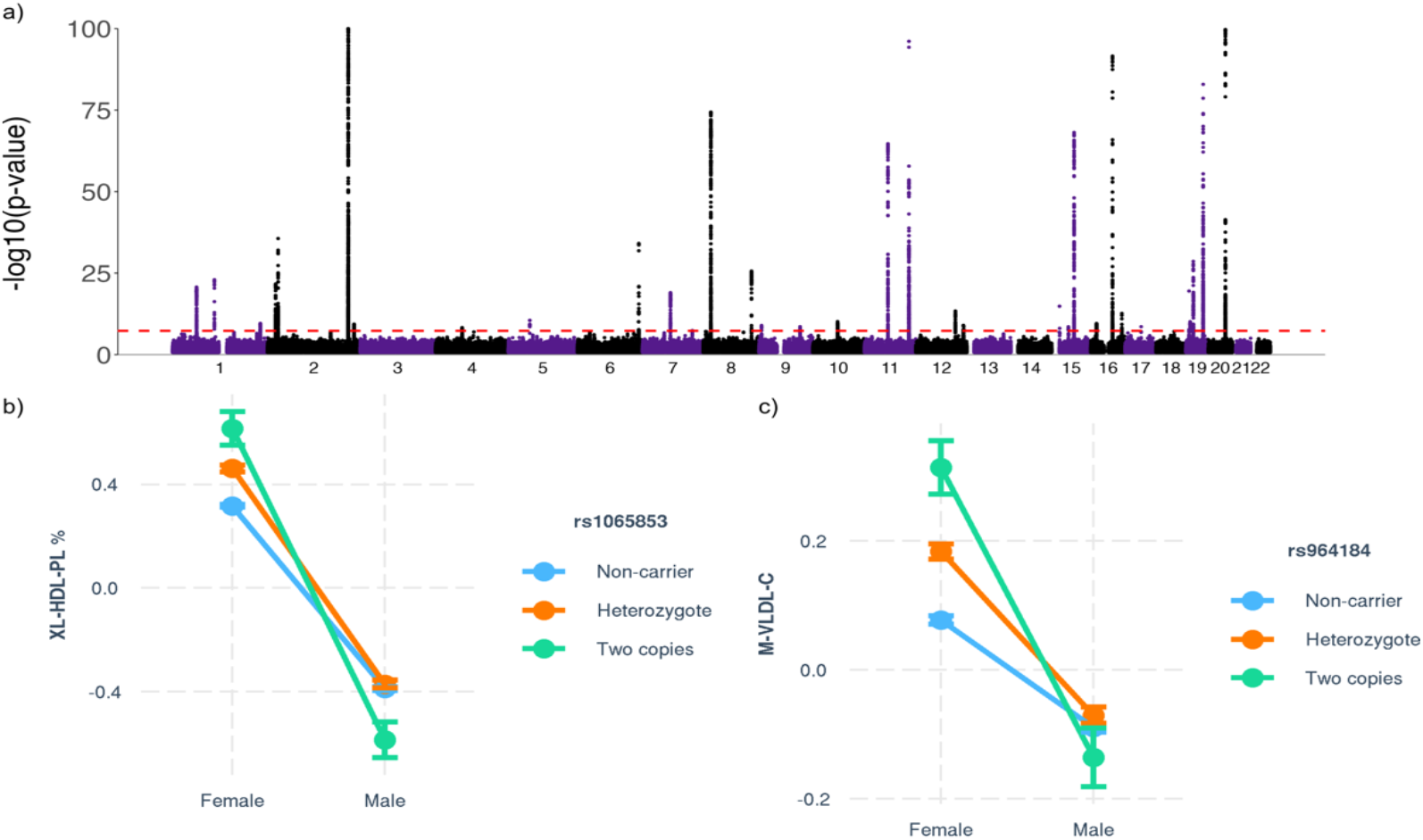
Genome-wide interactions between sex and genetic variants. **a)** Manhattan plot of the multivariate GWAS with an interaction term with sex on all 249 metabolites, with the observed -log10(p) of each interaction shown on the y-axis. The x-axis shows the relative genomic location, grouped by chromosome, and the red dashed line indicates the genome-wide significance threshold of 5×10^−8^. The y-axis is clipped at - log10(p)=150. **b)** Illustration of an identified significant cross-over interaction between sex and rs1065853 on chromosome 19, showing opposite effects on phospholipid concentrations (y-axis) in men and women (x-axis). **c)** An interaction effect of rs964184 on chromosome 11, illustrating effects on VLDL cholesterol concentrations only in women.

The concordance rate was 90.7% in the non-White UKB subset, with 268 out of the 496 lead variants being nominal significant (54.0%). In EstBB, the concordance rate was 99.3%, and 113 out of 158 (71.5%) of the available lead variants was nominally significant. The lists of all multivariate and univariate loci with significant interactions are provided in Supplementary Table 13.

Functional annotation of 29 genes mapped to the 31 MOSTest-identified locus lead variants through OpenTargets revealed tissue-specific upregulation in kidney, liver, and heart tissues based on GTEx v8 data, and enrichment for GO pathways involved primarily in cholesterol regulation. Coupling these 29 genes to the GWAS Catalog showed enrichment among gene lists reported for metabolic syndrome, CAD, T2D, and steatotic liver disease, which are well-known for having sex differences in prevalence and etiology.^12^.

Next, to estimate the causal nature of the identified relationships between BMI and metabolites, we ran bidirectional two-sample Mendelian randomization (MR), combining inverse variance weighted MR with the weighted median approach. There were no instances where the metabolites had a significant causal effect on BMI consistently across the different MR methods. BMI had a multiple comparisons-corrected significant causal effect on 79 metabolites, consistent across both MR methods, with strong negative effects on HDL cholesterol metabolites and positive effects on several of the amino acids, as summarized in **Figure 7**. When further thresholded by the MR Egger approach, the causal effect of BMI on only six metabolites remained: albumin, phenylalanine, average diameter for LDL particles, cholesterol % in small LDL, tyrosine, and valine.

**Figure 7.**
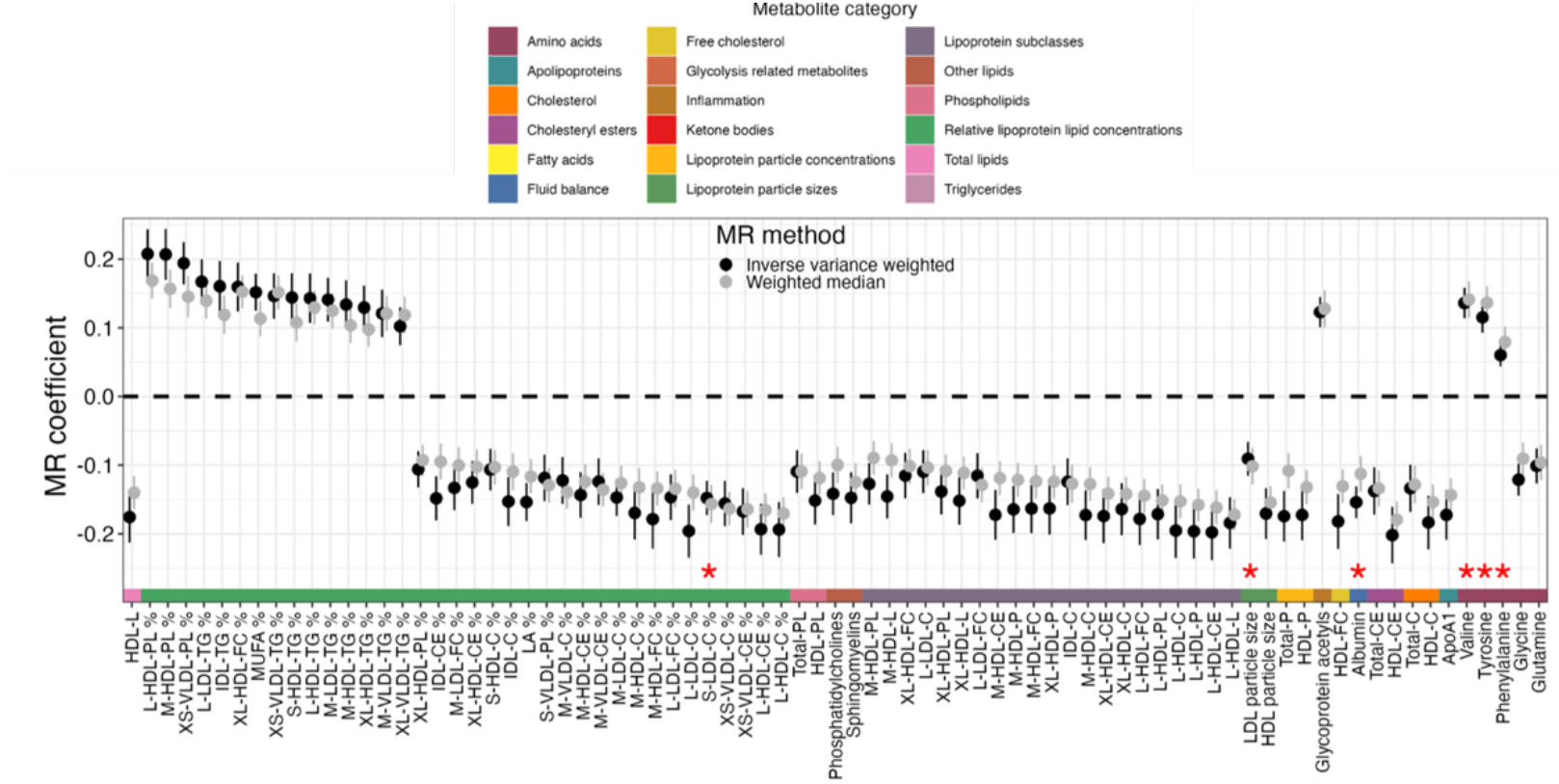
Causal influences of BMI on the metabolites. Plot listing coefficients from Mendelian Randomization (MR) analyses on the x-axis, and the 79 different metabolites that showed a significant influence of BMI on the y-axis. The dots and lines represent the point estimates with their standard errors, colour-coded by the MR method used, grouped by metabolite category, as specified in the legend above. The red stars indicate the 6 six metabolites that also show a significant causal effect of BMI using the MR-Egger method.

### Sensitivity analyses

We ran two sets of variations on the primary GWAS, to investigate the role of medication and of the preprocessing pipeline. First, we re-ran the primary GWAS controlling for insulin, blood pressure, and cholesterol-lowering medication. Second, we re-ran without the ‘ukbnmr’ pre-processing pipeline, directly on the originally released metabolomics data. For both variations, the produced summary statistics were highly comparable with the primary GWAS, with median genetic correlations of .992 and .998 across the metabolites, respectively.

## Discussion

Here, we reported results from a large-scale GWAS of circulating metabolite concentrations. This led to the identification of the largest number of discovered genetic determinants across these metabolites to date, mapped to genes with roles in lipid homeostasis. Our findings emphasized the pervasive pleiotropy across metabolic measures, with a sizeable role for rare variants. We further identified the causal effect of BMI on metabolites, indicating obesity as a primary target for improving metabolic health. Last, we discovered novel, sex-specific genetic effects on metabolite concentrations, which may explain the substantial sex differences in metabolic health.

Locus discovery was high, in line with the estimated genetic architecture. The complementary univariate and multivariate GWAS approaches employed in this study particularly emphasized the pervasive pleiotropy across the set of included metabolites, in accordance with previous findings.^6^ Joint analyses of these interrelated measures are essential to boost discovery of variants with small, yet distributed effects. The clinical relevance of this discovery is underscored by the results of the pheWAS analyses, showing the association of many of these pleiotropic variants with medical conditions across domains. This likely contributes to the extensive comorbidity across complex medical conditions with a cardiometabolic component,^28,29^ which is an important determinant of clinical outcomes.^29,30^

The gene-based analyses illustrated the relative contributions of common and rare variation, with extensive pleiotropy, to determining metabolite levels. The WES gene burden tests, aggregating across rare variants, identified 335 genes with widespread associations across both lipid and non-lipid metabolite categories. Among the most pleiotropic were apolipoprotein genes, well-known for their involvement in diabetes and CAD as well as in brain disorders.^31^ Particularly notable in this context is the identification of *BACE1* on chromosome 11 among the most influenced genes, the protein product of which is central to the generation of amyloid-B peptides in neurons and a key enzyme in the pathophysiology of AD.^32^ Overall, this rare variant data confirms the presence of impactful rare variants with high potential for druggability, as confirmed by the coupling to DGIdb. The generated data on the specificity of these genetic effects on metabolites is important information for research into comorbidities and for predicting utility as a biomarker and drug target.

The findings of the gene-by-sex interaction analyses underscore the substantial differences between males and females in metabolism.^11^ This is likely to be a strong explanatory factor of sex differences in the prevalence of a wide array of cardiometabolic conditions,^12^ advocating for the investigation of sex-specific mechanisms. The notoriously low power of interaction effects^33^ is counteracted by our multivariate approach. MOSTest is insensitive to differences in the directions of these interactions across the univariate measures, which would hamper other approaches to aggregation across measures. The identification of the widespread sex-dependent effects of rs1065853 showcases the potential of these interaction terms to identify variants that explain interindividual variation beyond their main effects. This SNP, located in a known enhancer of *APOE*, is well known for its association with numerous metabolic and clinical outcomes, including AD and CAD.^34,35^ The identification of such non-linear effects represent a new frontier in genomics, which needs to be explored in order to further resolve interindividual heterogeneity. Our findings particularly suggest value of additional sex-specific research into obesity and metabolic health.

The MR analyses provided evidence for the causal effect of BMI, as a proxy of obesity, on circulating metabolic biomarkers, emphasizing the importance of obesity as a primary target for treatment of cardiometabolic conditions. In accordance with previous findings in smaller samples, we show that BMI has a significant causal effect on levels of metabolites,^36^ while there was no evidence of effects of metabolites on BMI. Obesity therefore appears to drive changes in metabolites, which may then cause complications.^37^ Robustness analyses, sensitive to violations of MR assumptions,^38^ did substantially reduce the number of causal relationships identified. This suggests a sizeable role for pleiotropic effects complicating the relationship between genetically mediated obesity and metabolite levels, particularly the lipid-related measures, in line with our GWAS findings. It speaks, for instance, to the complex role of the GLP-1 secretory system, currently hailed as a highly promising therapeutic target for treatment of obesity,^39^ with divergent findings across both human and animal studies.^40^ A better understanding of the role of genetic susceptibility and sources of interindividual variation is needed to optimize individual outcomes.

Strengths of this study are the large sample size and the use of high quality, accurately measured metabolomics data. We further combined the study of individual metabolites with a multivariate approach to genetic discovery and inclusion of WES data, allowing for greater insight into the overall genetic architecture of metabolism. This study included two replication samples with varying genetic ancestry, enabling estimation of generalization of the findings. However, given known ethnic differences in the association between obesity and metabolic conditions such as T2D,^41^ the role of ethnicity should be investigated in further detail. It should also be noted that the data collection was not done under fasting conditions, which has been shown to obscure associations between genetic variation and metabolites.^19^

To conclude, metabolic health is central to the most prevalent and impactful medical conditions in our society, indicating a strong need for new therapeutic targets. Knowledge about causal individual-level determinants is central to develop effective strategies that optimally treat the individual. Here, we showed that accurate NMR-derived circulating metabolite concentrations share genetic influences that can be leveraged to boost discovery of pleiotropic variants of high relevance for cardiometabolic diseases. The summary statistics made freely available can be used by follow-up studies to further enhance our understanding of metabolism and related diseases, identify potential drug targets for these diseases, and contribute to the development of precision medicine by identifying individual-level determinants.

## Data Availability

The data incorporated in this work were gathered from public resources. All data described are available through UK Biobank, subject to approval from the UK Biobank access committee. See https://www.ukbiobank.ac.uk/enable-your-research/apply-for-access for further details. The MOSTest code is available via https://github.com/precimed/mostest (GPLv3 license). GWAS summary statistics are uploaded to the NHGRI-EBI GWAS catalog (https://www.ebi.ac.uk/gwas/).

## Methods

### Participants

For the UKB, we obtained data under accession number 27412. The composition, set-up, and data gathering protocols of the UKB have been extensively described elsewhere.^42^ It has received ethics approval from the National Health Service National Research Ethics Service (ref 11/NW/0382), and obtained informed consent from its participants. For the primary analyses, we selected unrelated White Europeans (KING cut-off 0.05)^43^ that had the Nightingale metabolomics data, as well as genetic and complete covariate data available (N=207 836, mean age 57.4 years (SD=8.0), 53.7 % female). BMI was taken from UKB field 21001, with a mean of 27.4 (SD=4.8). For the generalization analyses, we made use of data from non-White European UKB participants (N=27 509, mean age 54.5 years (SD=8.4), 54.3 % female). Ethnicity was based on self-report confirmed by genetics (UKB field 22006).

EstBB is a volunteer-based biobank composed of ∼213,000 individuals with data available on genotype, phenotype and electronic health records.^44^ All EstBB participants have signed an informed consent form and the study was granted ethical approval from the Estonian Council on Bioethics and Human Research (24 March 2020, nr 1.1-12/624). All analyses were conducted using data according to release S60 from EstBB. Specifically, individuals were selected under conditions identical to those used for the UKB data for filtering and quality control, resulting in 92,661 unrelated White European participants, with a mean age of 50.9 years (SD=16.2 years), 65.7% female. BMI values (mean 26.1, SD=5.3) were either calculated at the time of recruitment and blood donation or referenced from EHR within a year from enrollment.

### Data collection and pre-processing

We included all 249 metabolites from the Nightingale NMR metabolomics panel, encompassing 228 lipids, lipoproteins or fatty acids and 21 non-lipid traits, namely amino acids, ketone bodies, fluid balance, glycolysis-, and inflammation-related metabolites, as QC’ed and released by UKB.^2^ We applied additional pre-processing through the ‘ukbnmr’ R package, to remove sources of technical noise.^45^

We applied rank-based inverse normal transformation^46^ to each measure, leading to normally distributed measures as input for the GWAS.

### Univariate GWAS and univariate interaction GWAS

We made use of the UKB v3 imputed data, which has undergone extensive quality control procedures as described by the UKB genetics team.^47^ After converting the BGEN format to PLINK binary format,^48^ we set a minor allele frequency threshold of 0.005, leaving 11,144,506 SNPs.

We carried out univariate GWAS on each of the 249 metabolites through PLINK2, which were then combined into a multivariate GWAS through the freely available MOSTest software (https://github.com/precimed/mostest). Details about the procedure and its extensive validation have been described previously.^7^ GWAS on each of the normalized measures were carried out using the standard additive model of linear association between genotype vector, *g*_*j*_, and phenotype vector, *y*. In all analyses we covaried for mean-centered age and twenty genetic principal components. We additionally covaried for biological sex, except in the sex-specific analyses.

Association of genotype*sex interaction with each of 249 metabolites was tested with PLINK2, including genotype, sex, mean-centered age and 20 genetic principal components as covariates. Produced univariate GWASs were then combined into multivariate MOSTest analysis. Calibration of the null distribution for the MOSTest analysis was performed permuting both genotypes and sex independently.

### Clumping

For both univariate and multivariate GWAS, independent significant variants and genomic loci were identified in accordance with the Psychiatric Genomics Consortium locus definition.^49^ First, we selected a subset of variants that passed genome-wide significance threshold, and used PLINK to perform a clumping procedure at LD r^2^=0.6 to identify the list of independent significant variants. Second, we queried the reference panel for all candidate variants in LD r^2^ of 0.1 or higher with any independent significant variant. Further, for each independent significant variant, its corresponding genomic loci were defined as a contiguous region of the independent significant variants’ chromosome, containing all candidate variants in r^2^=0.1 or higher LD with the independent significant variant. Adjacent genomic loci were merged if separated by less than 250 KB. A subset of independent significant variants with LD r2<0.1 was selected as lead variants (with potentially more than one lead variant per locus). Finally for each locus the most significant among all lead variants was defined as the locus lead variant. Allele LD correlations were computed from EUR population of the 1000 genomes Phase 3 data. The number of unique significant loci across all univariate GWAS was determined through the min-P approach.^50^

### Gene mapping

We used the Variant-to-Gene (V2G) pipeline from Open Targets Genetics, to map lead variants to genes based on the strongest evidence from quantitative trait loci (QTL) experiments, chromatin interaction experiments, in *silico* functional prediction, and proximity of each variant to the canonical transcription start site of genes.^23^

### PheWAS

We used the ‘otargen’ R package to conduct the pheWAS analyses on each of the 534 MOSTest-identified locus lead SNPs. We restricted the analyses to the FinnGen and GWAS Catalog study sources, and selected only traits that had the term ‘disease’ in the trait category. The results were thresholded to associations of each of the locus lead SNP at p<.05 divided by the unique number of traits included (n=7,684).

### Fine-mapping procedure

We used the SAFFARI pipeline to perform statistical and functional fine mapping.^22^ This consisted of applying PolyFun+FINEMAP to each of the GWAS in order to identify sets of functionally-informed highly credible causal variants, selecting those that were part of a credible set with a posterior probability >.95 prioritizing these for follow-up.

### WES gene burden tests

We used Regenie (v3.1.1) to perform omnibus SKAT-O tests to combine variance component tests and burden tests for each of the 249 metabolites, with age, sex and 20 genetic principal components as covariates. We merged the genotype data of chromosome 1 to 22 into a single PLINK file, lifted the genomic build from GRCh37 to GRCh38, and filtered with PLINK (--maf 0.01 --mac 20 --geno 0.1 --hwe 1e-15 --mind 0.1) to select 591,260 SNPs for step 1. Step 2 variants were rare (MAF < 0.005) with the following annotation masks: LoF, missense (0/5), missense (5/5), missense (>=1/5), and synonymous. We used relevant annotation files described elsewhere: https://biobank.ctsu.ox.ac.uk/crystal/refer.cgi?id=916.

We included the same set of protein coding genes as used for the MAGMA gene-based analyses. The analyses were conducted on the Research Analysis Platform. (https://ukbiobank.dnanexus.com)

### Gene-set analyses

We carried out gene-based analyses using MAGMA v1.08 with default settings, which entails the application of a SNP-wide mean model.^51^ We used a randomly selected set of 10 000 white British UKB participants as reference panel. Gene-set analyses were done in a similar manner, restricting the sets under investigation to those that are part of the Gene Ontology biological processes subset (n=7,522), as listed in the Molecular Signatures Database (MsigdB; c5.bp.v7.1).

For tissue-specificity analyses, we applied MAGMA gene-property analyses to test relationships between tissue-specific gene expression profiles and the identified gene associations. This encompassed running one-sided tests for each of 30 general tissue types, testing whether the association between each tissue’s known gene expression levels and the gene-based Z-scores is greater than 0, corrected for the average expression across all tissue types and a set of technical confounders. We used preprocessed and normalized GTEx v8 tissue expression values^52^ as provided through FUMA’s downloads (https://fuma.ctglab.nl/).

Multiple comparison’s correction for these analyses consisted of a Bonferroni correction for the number of protein-coding genes, with α=.05/17,849=2.8*10^−6^

### Drug enrichment analysis

The Drug Gene Interaction Database (DGIdb, (https://www.dgidb.org/) v.5.0.6 (04/04/2024)^25^ was used to identify drug-gene interactions among the genes identified from the WES gene burden tests. The DGIdb provides information on drug-gene interactions from 28 diverse sources that are aggregated and normalized. The database collects drug-gene interactions based on information about therapeutic targets and their corresponding drugs, knowledge from clinical trials, as well as potentially clinically actionable drug-gene associations based on metadata such as molecule structure and molecular weight.^25^ Gene-set enrichment analysis (GSEA) was performed to test if the genes identified from the WES gene burden tests were significantly (FDR<0.05) enriched for targets of specific drugs.

### LDSC

We applied univariate^53^ and cross-trait^54^ LDSC to estimate narrow-sense heritability and genetic correlations, respectively. For this, we formatted the GWAS summary statistics using our standardized pipeline, including ‘munging’ and removal of all variants in the extended major histocompatibility complex (MHC) region (chr6:25–35 Mb), in accordance with recommendations (https://github.com/precimed/python_convert/blob/master/sumstats.py).

### MiXeR analysis

We applied a causal mixture model^15,16^ to estimate the percentage of variance explained by genome-wide significant SNPs as a function of sample size. For each SNP, *i*, MiXeR models its additive genetic effect of allele substitution,*β*_*i*_, as a point-normal mixture, 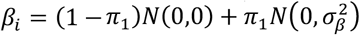, where *π*_1_ represents the proportion of non-null SNPs (‘polygenicity’) and 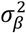 represents the variance of effect sizes of non-null SNPs (‘discoverability’). Then, for each SNP, *j*, MiXeR incorporates LD information and allele frequencies for 9,997,231 SNPs extracted from the EUR population of the 1000 Genomes Phase3 data to estimate the expected probability distribution of the signed test statistic, 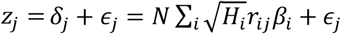, where *N* is the sample size, *H*_*i*_ indicates heterozygosity of i-th SNP, *r*_*ij*_ indicates an allelic correlation between i-th and j-th SNPs, and 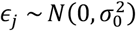 is the residual variance. Further, the three parameters, *π*_1_, 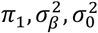, are fitted by direct maximization of the likelihood function. Finally, given the estimated parameters of the model, the power curve *S*(*N*) is then calculated from the posterior distribution *p*(*δ*_*j*_|*z*_*j*_, *N*.

For quality control of the MiXeR results, we used the Aikaike Information Criterion (AIC), comparing the Gaussian mixture model fit to that of the infinitesimal model. In this study, the AIC values of all 249 metabolites were positive, i.e. the Gaussian mixture had better model fit, warranting the inclusion of the results.

### Mendelian randomization

We ran bidirectional MR, investigating the causal relationships between BMI and the 249 metabolites, with the *TwoSampleMR* R package. For this, we combined the BMI GWAS summary statistics from the GIANT consortium with no UKB participants (N=339 224),^55^ to prevent sample overlap, with the metabolomics GWAS summary statistics generated in this study. We selected only genome wide significant variants for the analysis, clumped using PLINK with clump_p = 1, clump_r2 = 0.001, clump_kb = 10000 against the 1000 Genomes Phase3 503 EUR samples keeping other settings default. We calculated MR regression coefficients using the inverse variance weighted method and the weighted median method. To create robust findings, we only selected findings that showed a multiple comparisons-significance (p<.05/249) across both methods. As an additional check, we ran MR-Egger and selected those relationships with nominal significance on this test.

### Statistical analyses

All pre-processing steps and analyses performed outside the above-mentioned tools and software, e.g. formatting the data and creating the graphs, were carried out in R, v4.2.

## Acknowledgements

This work was partly performed on the TSD (Tjeneste for Sensitive Data) facilities, owned by the University of Oslo, operated and developed by the TSD service group at the University of Oslo, IT-Department (USIT). Computations were also performed on resources provided by UNINETT Sigma2 - the National Infrastructure for High-Performance Computing and Data Storage in Norway. Computation of Estonian Biobank data was carried out in part in the High-Performance Computing Center of University of Tartu. The Estonian Biobank Research Team, responsible for data collection, genotyping, QC, and imputation, comprises Andres Metspalu, Tõnu Esko, Reedik Mägi, Mari Nelis and Georgi Hudjashov.

## Conflicts of interest

OAA has received speaker fees from Lundbeck, Janssen, Otsuka, and Sunovion and is a consultant to Cortechs.ai. and Precision Health. AMD was a Founder of and holds equity in CorTechs Labs, Inc, and serves on its Scientific Advisory Board. He is also a member of the Scientific Advisory Board of Human Longevity, Inc. (HLI), and the Mohn Medical Imaging and Visualization Centre in Bergen, Norway. He receives funding through a research agreement with General Electric Healthcare (GEHC). The terms of these arrangements have been reviewed and approved by the University of California, San Diego in accordance with its conflict-of-interest policies. OF is a consultant to Precision Health All other authors report no potential conflicts of interest.

## Financial support

The authors were funded by the Research Council of Norway (273291, 273446, 296030, 324252, 324499, 326813), NordForsk (164218), the South-Eastern Norway Regional Health Authority 2017-112., Estonian Research Council (grant PSG615), The University of Tartu grant PLTGI24925, European Union’s Horizon 2020 research and innovation program under grant agreement (CoMorMent) 847776, 964874 (RealMent).

## Author contributions

D.v.d.M., Z.R., A.S., S.D., and O.A.A. conceived the study; D.v.d.M., Z.R., A.O., G.K., and A.S. pre-processed the data. D.v.d.M., Z.R. A.O., E.M.K. and A.S. performed all analyses, with conceptual input from J.R., S.S., P.P., S.D. and O.A.A.; All authors contributed to interpretation of results; D.v.d.M. drafted the manuscript and all authors contributed to and approved the final manuscript.

---

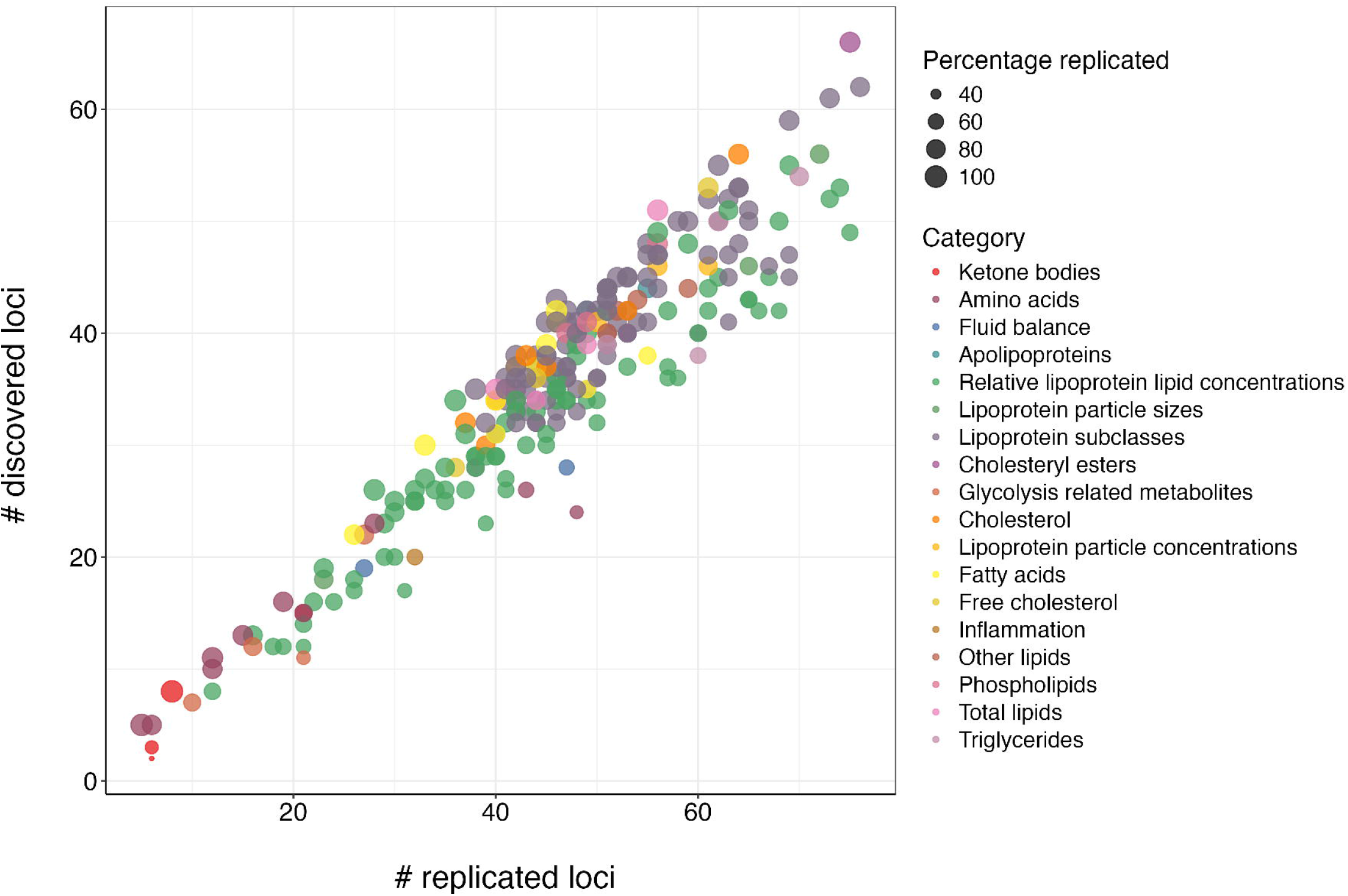

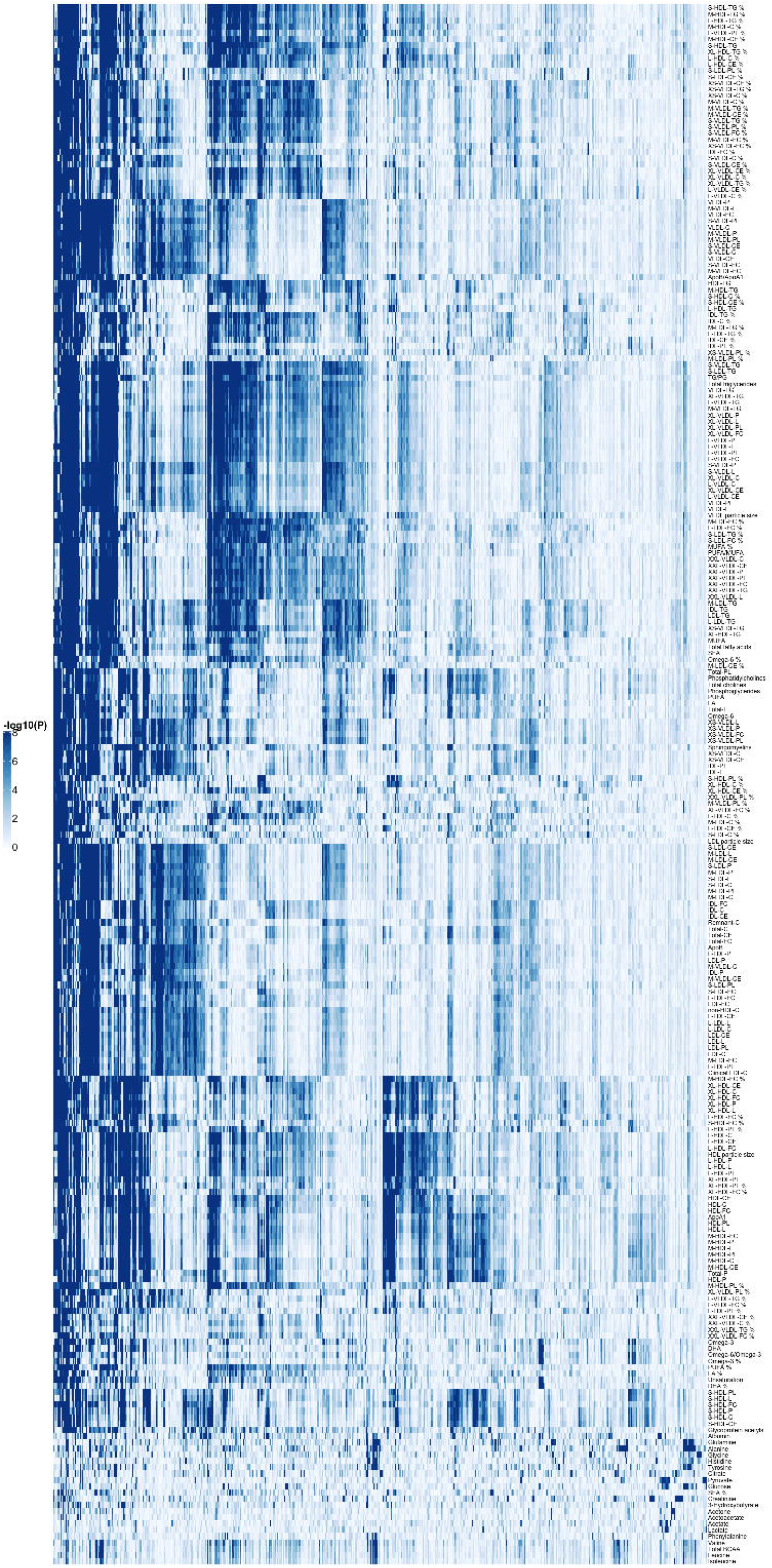

## Notes

### Author Declarations

UKB has received ethics approval from the National Health Service National Research Ethics Service (ref 11/NW/0382) and obtained informed consent from its participants. All EstBB participants have signed an informed consent form and the study was granted ethical approval from the Estonian Council on Bioethics and Human Research (24 March 2020, nr 1.1-12/624).

## References

1. Würtz, P. et al. Quantitative Serum Nuclear Magnetic Resonance Metabolomics in Large-Scale Epidemiology: A Primer on-Omic Technologies. Am. J. Epidemiol. 186, 1084–1096 (2017).

2. Julkunen, H. et al. Atlas of plasma NMR biomarkers for health and disease in 118,461 individuals from the UK Biobank. Nat. Commun. 14, 604–604 (2023).

3. Nightingale Health Biobank Collaborative Group et al. Metabolomic and genomic prediction of common diseases in 477,706 participants in three national biobanks. medRxiv 2023.06.09.23291213 (2023) doi:10.1101/2023.06.09.23291213.

4. Saklayen, M. G. The global epidemic of the metabolic syndrome. Curr. Hypertens. Rep. 20, 1–8 (2018).

5. Karsten |. A Table of all Published GWAS with metabolomics. Human Metabolic Individuality http://www.metabolomix.com/list-of-all-published-gwas-with-metabolomics/ (2024).

6. Smith, C. J. et al. Integrative analysis of metabolite GWAS illuminates the molecular basis of pleiotropy and genetic correlation. Elife 11, e79348 (2022).

7. van der Meer, D. et al. Understanding the genetic determinants of the brain with MOSTest. Nat. Commun. 11, 3512 (2020).

8. Bomba, L. et al. Whole-exome sequencing identifies rare genetic variants associated with human plasma metabolites. Am. J. Hum. Genet. 109, 1038–1054 (2022).

9. Abhishek Nag et al. Assessing the contribution of rare-to-common protein-coding variants to circulating metabolic biomarker levels via 412,394 UK Biobank exome sequences. medRxiv 2021.12.24.21268381 (2021) doi:10.1101/2021.12.24.21268381.

10. Park, S., Sadanala, K. C. & Kim, E.-K. A metabolomic approach to understanding the metabolic link between obesity and diabetes. Mol. Cells 38, 587–596 (2015).

11. Varlamov, O., Bethea, C. L. & Roberts Jr, C. T. Sex-specific differences in lipid and glucose metabolism. Front. Endocrinol. 5, 241 (2015).

12. Gerdts, E. & Regitz-Zagrosek, V. Sex differences in cardiometabolic disorders. Nat. Med. 25, 1657–1666 (2019).

13. Link, J. C. & Reue, K. Genetic Basis for Sex Differences in Obesity and Lipid Metabolism. Annu. Rev. Nutr. 37, 225–245 (2017).

14. Privé, F., Luu, K., Blum, M. G. B., McGrath, J. J. & Vilhjálmsson, B. J. Efficient toolkit implementing best practices for principal component analysis of population genetic data. Bioinformatics 36, 4449–4457 (2020).

15. Holland, D. et al. Beyond SNP heritability: Polygenicity and discoverability of phenotypes estimated with a univariate Gaussian mixture model. PLoS Genet. 16, e1008612 (2020).

16. Frei, O. et al. Bivariate causal mixture model quantifies polygenic overlap between complex traits beyond genetic correlation. Nat. Commun. 10, 2417 (2019).

17. van der Meer, D. et al. Quantifying the polygenic architecture of the human cerebral cortex: extensive genetic overlap between cortical thickness and surface area. Cereb. Cortex 30, 5597–5603 (2020).

18. Li, J. & Ji, L. Adjusting multiple testing in multilocus analyses using the eigenvalues of a correlation matrix. Heredity 95, 221–227 (2005).

19. Karjalainen, M. K. et al. Genome-wide characterization of circulating metabolic biomarkers. Nature (2024) doi:10.1038/s41586-024-07148-y.

20. Pendergrass, S. et al. The use of phenome-wide association studies (PheWAS) for exploration of novel genotype-phenotype relationships and pleiotropy discovery. Genet. Epidemiol. 35, 410–422 (2011).

21. Penninx, B. W. & Lange, S. M. Metabolic syndrome in psychiatric patients: overview, mechanisms, and implications. Dialogues Clin. Neurosci. 20, 63–73 (2018).

22. Maria Koromina et al. Fine-mapping genomic loci refines bipolar disorder risk genes. medRxiv 2024.02.12.24302716 (2024) doi:10.1101/2024.02.12.24302716.

23. Mountjoy, E. et al. An open approach to systematically prioritize causal variants and genes at all Published human GWAS trait-associated loci. Nat. Genet. 53, 1527–1533 (2021).

24. Lee, S. et al. Optimal unified approach for rare-variant association testing with application to small-sample case-control whole-exome sequencing studies. Am. J. Hum. Genet. 91, 224–237 (2012).

25. Cannon, M. et al. DGIdb 5.0: rebuilding the drug–gene interaction database for precision medicine and drug discovery platforms. Nucleic Acids Res. 52, D1227–D1235 (2024).

26. Alcala-Diaz, J. F. et al. A gene variation at the ZPR1 locus (rs964184) interacts with the type of diet to modulate postprandial triglycerides in patients with coronary artery disease: from the coronary diet intervention with olive oil and cardiovascular prevention study. Front. Nutr. 9, 885256 (2022).

27. Ueyama, C. et al. Association of FURIN and ZPR1 polymorphisms with metabolic syndrome. Biomed. Rep. 3, 641–647 (2015).

28. Kivimäki, M. et al. Overweight, obesity, and risk of cardiometabolic multimorbidity: pooled analysis of individual-level data for 120 813 adults from 16 cohort studies from the USA and Europe. Lancet Public Health 2, e277–e285 (2017).

29. Correll, C. U. et al. Cardiometabolic comorbidities, readmission, and costs in schizophrenia and bipolar disorder: a real-world analysis. Ann. Gen. Psychiatry 16, 1–8 (2017).

30. Canoy, D. et al. Association between cardiometabolic disease multimorbidity and all-cause mortality in 2 million women and men registered in UK general practices. BMC Med. 19, 1–10 (2021).

31. Elliott, D. A., Weickert, C. S. & Garner, B. Apolipoproteins in the brain: implications for neurological and psychiatric disorders. Clin. Lipidol. 5, 555–573 (2010).

32. Cole, S. L. & Vassar, R. The Alzheimer’s disease β-secretase enzyme, BACE1. Mol. Neurodegener. 2, 1–25 (2007).

33. Clayton, D. G. Prediction and interaction in complex disease genetics: experience in type 1 diabetes. PLoS Genet. 5, e1000540– e1000540 (2009).

34. Strawbridge, R. J. et al. Carotid intima-media thickness: novel loci, sex-specific effects, and genetic correlations with obesity and glucometabolic traits in UK Biobank. Arterioscler. Thromb. Vasc. Biol. 40, 446–461 (2020).

35. Lutz, M. W. & Chiba-Falek, O. Bioinformatics pipeline to guide late-onset Alzheimer’s disease (LOAD) post-GWAS studies: Prioritizing transcription regulatory variants within LOAD-associated regions. Alzheimers Dement. Transl. Res. Clin. Interv. 8, e12244 (2022).

36. Bell, J. A. et al. Effects of general and central adiposity on circulating lipoprotein, lipid, and metabolite levels in UK Biobank: a multivariable Mendelian randomization study. Lancet Reg. Heal. 21, (2022).

37. Marott, S. C., Nordestgaard, B. G., Tybjærg-Hansen, A. & Benn, M. Causal associations in type 2 diabetes development. J. Clin. Endocrinol. Metab. 104, 1313–1324 (2019).

38. Burgess, S. & Thompson, S. G. Interpreting findings from Mendelian randomization using the MR-Egger method. Eur. J. Epidemiol. 32, 377–389 (2017).

39. Drucker, D. J. GLP-1 physiology informs the pharmacotherapy of obesity. Mol. Metab. 57, 101351 (2022).

40. Hira, T., Pinyo, J. & Hara, H. What is GLP-1 really doing in obesity? Trends Endocrinol. Metab. 31, 71–80 (2020).

41. Chiu, M., Austin, P. C., Manuel, D. G., Shah, B. R. & Tu, J. V. Deriving ethnic-specific BMI cutoff points for assessing diabetes risk. Diabetes Care 34, 1741–1748 (2011).

42. Sudlow, C. et al. UK biobank: an open access resource for identifying the causes of a wide range of complex diseases of middle and old age. PLoS Med. 12, e1001779–e1001779 (2015).

43. Manichaikul, A. et al. Robust relationship inference in genome-wide association studies. Bioinformatics 26, 2867–2873 (2010).

44. Leitsalu, L. et al. Cohort profile: Estonian biobank of the Estonian genome center, university of Tartu. Int. J. Epidemiol. 44, 1137–1147 (2015).

45. Ritchie, S. C. et al. Quality control and removal of technical variation of NMR metabolic biomarker data in ∼120,000 UK Biobank participants. Sci. Data 10, 64–64 (2023).

46. Beasley, T. M., Erickson, S. & Allison, D. B. Rank-based inverse normal transformations are increasingly used, but are they merited? Behav. Genet. 39, 580–595 (2009).

47. Bycroft, C. et al. The UK Biobank resource with deep phenotyping and genomic data. Nature 562, 203–209 (2018).

48. Chang, C. C. et al. Second-generation PLINK: rising to the challenge of larger and richer datasets. Gigascience 4, 7–7 (2015).

49. Watanabe, K., Taskesen, E., Bochoven, A. & Posthuma, D. Functional mapping and annotation of genetic associations with FUMA. Nat. Commun. 8, 1826–1826 (2017).

50. Van der Sluis, S., Posthuma, D. & Dolan, C. V. TATES: efficient multivariate genotype-phenotype analysis for genome-wide association studies. PLoS Genet. 9, e1003235–e1003235 (2013).

51. de Leeuw, C. A., Mooij, J. M., Heskes, T. & Posthuma, D. MAGMA: generalized gene-set analysis of GWAS data. PLoS Comput. Biol. 11, e1004219–e1004219 (2015).

52. Consortium, Gte. et al. The Genotype-Tissue Expression (GTEx) pilot analysis: multitissue gene regulation in humans. Science 348, 648–660 (2015).

53. Bulik-Sullivan, B. K. et al. LD Score regression distinguishes confounding from polygenicity in genome-wide association studies. Nat. Genet. 47, 291–295 (2015).

54. Bulik-Sullivan, B. et al. An atlas of genetic correlations across human diseases and traits. Nat. Genet. 47, 1236–1236 (2015).

55. Locke, A. E. et al. Genetic studies of body mass index yield new insights for obesity biology. Nature 518, 197–206 (2015).

